# MiR-139-5p is a causal biomarker for type 2 diabetes; Results from genome-wide microRNA profiling and Mendelian randomization analysis in a population-based study

**DOI:** 10.1101/2021.05.13.21257090

**Authors:** Michelle M.J. Mens, Rima Mustafa, Fariba Ahmadizar, M. Arfan Ikram, Marina Evangelou, Maryam Kavousi, Abbas Dehghan, Mohsen Ghanbari

## Abstract

MicroRNAs (miRNAs) have emerged as key regulators of gene expression. Differential expression of miRNAs has been linked to diabetes, but underlying pathways remain poorly understood. We performed genome-wide miRNAs profiling and tested the causal associations between miRNAs and type 2 diabetes in the general population. Subsequently, we investigated target genes and metabolites of miRNAs to provide insight into the metabolic disturbances that emerge with type 2 diabetes. Between 2002 and 2005, plasma levels of 2083 circulatory miRNAs were profiled in 1900 participants (mean age 71.4 years) of the population-based Rotterdam Study cohort. The associations of 591 well-expressed miRNAs with prevalent and incident type 2 diabetes were examined until 2015. Two-sample Mendelian Randomization (MR) was conducted to investigate the causal associations and miRNA-target genes and metabolites were studied in relation to type 2 diabetes. At baseline, higher plasma levels of miR-139-5p and miR-193a-5p were associated (FDR<0.05) with prevalent type 2 diabetes (n=253 cases). During a follow-up of >9.0 years, 209 participants developed type 2 diabetes. Plasma levels of miR-99a-5p, miR-4664-3p, miR-29a-3p, miR-122-5p, and miR-125b-5p were significantly associated with incident type 2 diabetes (n=209). Two-sample MR confirmed a causal effect for miR-139-5p (MR-IWV-beta=0.10, p=3.51×10^−4^) on type 2 diabetes. We found several target genes and metabolites that could link miR-139-5p to pathways underlying type 2 diabetes. Our study indicates a causal relationship between miR-139-5p and type 2 diabetes and suggests this miRNA as a plasma biomarker of type 2 diabetes.

## Introduction

MicroRNAs (miRNAs) are small non-coding RNA molecules that have been recognized as the fine-tuners of gene expression, through repressing gene transcription or degradation of messenger (m)RNAs. Previous studies have shown that miRNA expression levels, including levels of miR-126 (1) and miR-222 (2), vary between type 2 diabetes patients and non-patients. Recent advantages in high-throughput technologies as next-generations sequencing methods have made it possible to investigate a wide variety of miRNAs. This allows observational studies to agnostically test associations between miRNAs and complex diseases, like type 2 diabetes, in the general population. Yet previous studies were often subject to known epidemiologic biases including confounding and reverse causation. Increasing advantages of Mendelian randomization (MR) methods enable to infer causal associations between exposure and outcome using the random inheritance property of genetic variants, therefore, enable us to go beyond observational associations and assess causality (3).

Large scale research consortia studying genetic and metabolite markers have given us insight into underlying pathways involved in type 2 diabetes development (4, 5). Given the ability of miRNAs to target thousands of genes, miRNAs are likely also to play a regulatory role in the metabolic disturbance that causes type 2 diabetes. However, it remains unclear which specific miRNAs are the major players. In addition, the interaction between miRNAs, genes and metabolites with regard to type 2 diabetes has not yet been well described. In this study, we characterized 2083 miRNAs in an advanced population-based cohort and studied the associations of well-expressed miRNAs with prevalent and incident type 2 diabetes. Further, we studied the potential causal effect of the identified miRNAs on type 2 diabetes using MR approach and elaborate on the possible underlying pathways by investigating putative target genes and metabolites of these miRNAs.

## Methods

### Study Design and Participants

This study was embedded within the Rotterdam Study, a prospective population-based cohort among middle-aged and elderly participants in the suburb Ommoord in Rotterdam, the Netherlands. In 1990, 7,983 inhabitants aged 55 years or older were recruited to participate in the first cohort of the Rotterdam Study (RS-I) (78% response rate of 10,215 invitees). In 2000, the Rotterdam Study was extended by 3,011 participants that moved to Ommoord or turned 55 years old (RS-II). A detailed description of the Rotterdam Study can be found elsewhere (6). In the current study, genome-wide miRNA profiling was performed in a random subset (n=1000) of the fourth visit of Rotterdam Study-I (RS-I-4) and a random subset (n=1000) of the second visit of Rotterdam Study-II (RS-II-2). These visits were performed between 2002 and 2005 with follow-up visits every 4-5 years. From 2000 participants with miRNA data, we excluded participants with missing data on type 2 diabetes at baseline (n=99) and one participant because of missing data for all miRNAs. For the longitudinal study with incident type 2 diabetes, we additionally excluded prevalent type 2 diabetes cases at baseline (n=253).

The Rotterdam Study has been approved by the Medical Ethics Committee of the Erasmus MC (registration number MEC 02.1015) and by the Dutch Ministry of Health, Welfare, and Sport (Population Screening Act WBO, license number 1071272-159521-PG). The Rotterdam Study has been entered into the Netherlands National Trial Register (NTR; www.trialregister.nl) and into the WHO International Clinical Trials Registry Platform (ICTRP; www.who.int/ictrp/network/primary/en/) under shared catalog number NTR6831.

### Measurements and Definitions

Full details of the Rotterdam Study miRNA expression profiling and quality control have been described previously (7). In brief, plasma miRNA levels were determined using the HTG EdgeSeq miRNA Whole Transcriptome Assay (WTA), a next-generation sequencing (NGS)-based application that measures the expression of 2083 human miRNAs (HTG Molecular Diagnostics, Tuscon, AZ, USA), by the Illumina NextSeq 500 sequencer (Illumina, San Diego, CA, USA). Quantification of miRNA expression was based on counts per million (CPM). Log2 transformation of CPM was used as standardization and adjusted for total reads within each sample. A lower limit of quantification method was used for the normalization and selection of 591 well-expressed miRNAs that were used in the current analysis.

At baseline and during follow-up, type 2 diabetes cases were ascertained by the use of general practitioners’ records, hospital discharge letters, and serum glucose measurements collected from center visits. Glucose measurements were obtained during visits to the research center. Type 2 diabetes was defined according to the World Health Organization definition as fasting glucose levels of ≥7.0 mmol/L, non-fasting glucose levels ≥11.1 mmol/L, or the use of glucose-lowering medication (8). The follow-up for incident type 2 diabetes is complete until January 1, 2015. Participants were followed from study entry until occurrence of type 2 diabetes, death, last health status update when they were known to be free of type 2 diabetes, or January 1, 2015, whichever came first.

### Statistical Analysis

We used logistic regression and Cox proportional hazards regression analyses to determine the association between the plasma levels of 591 well-expressed miRNAs with prevalent and incident type 2 diabetes respectively. Odds ratios (OR) and Hazard ratios (HR) with 95% confidence interval (CI) were calculated for each additional unit log2 CPM miRNA expression. The first model was adjusted for age, sex, and cohort (RS-I-4 and RS-II-2). The second model was additionally adjusted for smoking status, alcohol use, body-mass index, high-density lipoprotein, serum total cholesterol, and hypertension. To reduce the possible bias induced by missing values, multiple imputations on confounders was performed. Values were imputed with a maximum iteration number of 10 (N=25 imputations) using the Markov Chain Monte Carlo method, R package “*mice*” (9). The Benjamini-Hochberg method was used to compute the false discovery rate (FDR) (10). Significant associations were considered when FDR<0.05. Furthermore, the identified miRNAs were categorized in ‘high or low expressed’ based on their median expression values and cumulative hazard graphs were generated. Analyses were performed using R version 3.6.1 (The R Foundation for Statistical Computing, Vienna, Austria).

### Mendelian randomization analysis

To assess the causal relationship between the identified miRNAs and type 2 diabetes, we used the two-sample MR approach given the specific conditions illustrated in **Figure 1**. Genetic instruments for each identified miRNA were obtained by performing genome-wide association studies (GWAS) in the Rotterdam Study (n=1687) adjusting for age, sex, and population stratification using the first five principal components. Single-nucleotide polymorphisms (SNPs) with minor allele frequency >5% and imputation quality (Rsq) >0.7 were retained. We selected the instruments as SNPs associated with each exposure (p= <1.0×10^−5^) and F-statistics>10 to avoid the weak instrument bias. SNPs were clumped using a linkage-disequilibrium threshold of r^2^<0.1 to remove correlated SNPs. Genetic association estimates between the instruments and type 2 diabetes were extracted from GWAS summary results on type 2 diabetes (74124 cases and 824006 controls) (5). Inverse variance weighted method (IVW) was used to combine the effect estimates of the genetic instruments. In the presence of heterogeneity, sensitivity methods such as weighted median (WM), MR-Egger and MR-PRESSO were used (11-13). MR analysis was performed using the MRCIEU/TwoSampleMR package in R (14).

**Figure 1.**
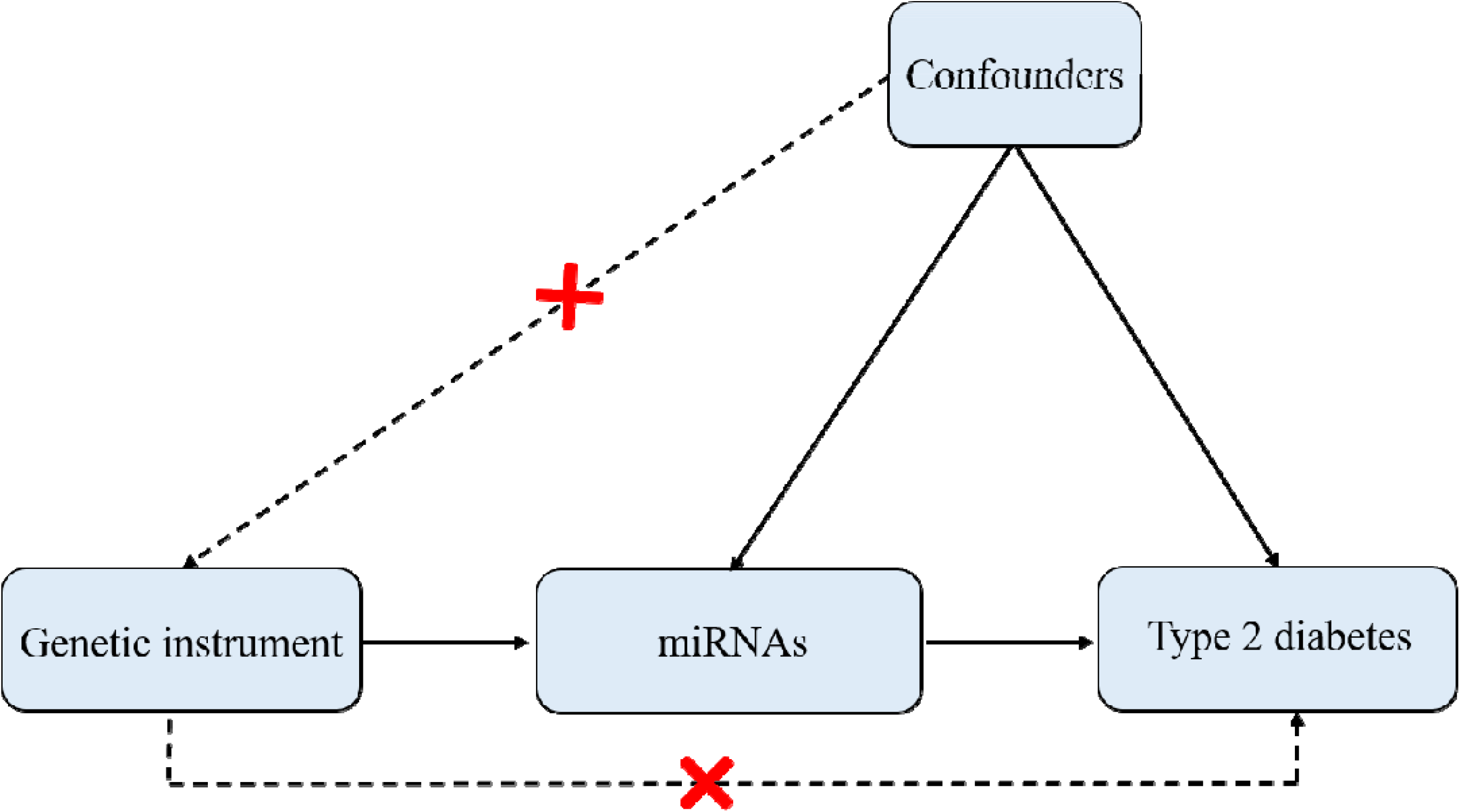
Overview of the MR process. Identified miRNAs at the observational approach were used as exposure. Type 2 diabetes was used as outcome.

### Post-hoc analysis

To further elucidate the underlying mechanisms involved in the pathology of type 2 diabetes, we tested if the genetic variants were also reported to be associated with metabolite levels using summary statistics of a cross-platform, genome-wide meta-analysis of 174 metabolites across six cohorts with different measurement platforms, consisting of 86507 participants (4). Subsequently, we performed a lookup to check if the genetic variants associated with the expression levels of miRNA of interest also explain the variation of gene expression (15).

To dissect the putative target genes of identified miRNAs, three commonly used miRNA target prediction databases, miRTarBase (16), TargetScan (v7.2) (17) and miRDB(18), were used. We chose to include putative target genes available in either two out of three databases or reported as validated target genes by strong validation methods, such as reporter assay, western blot, and qPCR. Then, we extracted SNPs in these target genes and tested the association with type 2 diabetes and fasting glucose using summary statistics data (5, 19).

## Results

Baseline characteristics of the study population are illustrated in **Table 1**. A total of 253 participants had prevalent type 2 diabetes at baseline and an additional 209 participants were diagnosed with incident diabetes during a mean follow-up period of 9.0 years (standard deviation (SD) 3.1). Across 591 tested miRNAs, miR-139-5p (OR 2.63; 95%CI 1.65-4.26; p=6.11×10^−5^) and miR-193a-5p (OR 1.91; 95%CI 1.42-2.56 p=1.86×10^−5^) were significantly associated with prevalent type 2 diabetes (FDR<0.05) after adjustments for covariates in model 2. In total, 51 miRNAs were nominally (p<0.05) associated with prevalent diabetes (**Figure 2A; Supplementary Table 1**). Furthermore, Cox proportional hazard regression analysis revealed four miRNAs significantly associated with an increased risk for incident diabetes. Among these were miR-99a-5p (HR 2.08; 95%CI 1.40-3.10; p=3.19×10^−4^), miR-29a-3p (HR 1.96; 95%CI 1.35-2.84; p=3.58×10^−4^), miR-122-5p (HR 1.29; 95%CI 1.12-1.49;p=3.65×10^−4^) and miR-125b-5p (HR 1.96; 95%CI 1.35-2.85; p=4.16×10^−4^). Moreover, we found miR-4664-3p significantly associated with a reduced risk for incident diabetes (HR 0.56; 95%CI 0.41-0.77; p=3.39×10^−4^). Summary statistics of the 77 nominally associated miRNAs for incident diabetes can be found in **Supplementary Table 2**. Notably, miR-139-5p and miR-193a-5p were nominally associated with incident diabetes (**Figure 2B**), but none of the five identified miRNAs for incident diabetes were nominally associated with prevalent diabetes. Also, the OR and HR of the seven identified miRNAs in each cohort (RS-I-4 and RS-II-2) are separately illustrated in **Supplementary Figure 1**. The observed associations were in a consistent direction between cohorts. Additionally, see **Supplementary Table 3** for the baseline characteristics split by median expression values of the seven miRNAs. Notably, glucose was significantly different in low versus high categorized expressed miRNAs of all seven miRNAs.

**Table 1.**
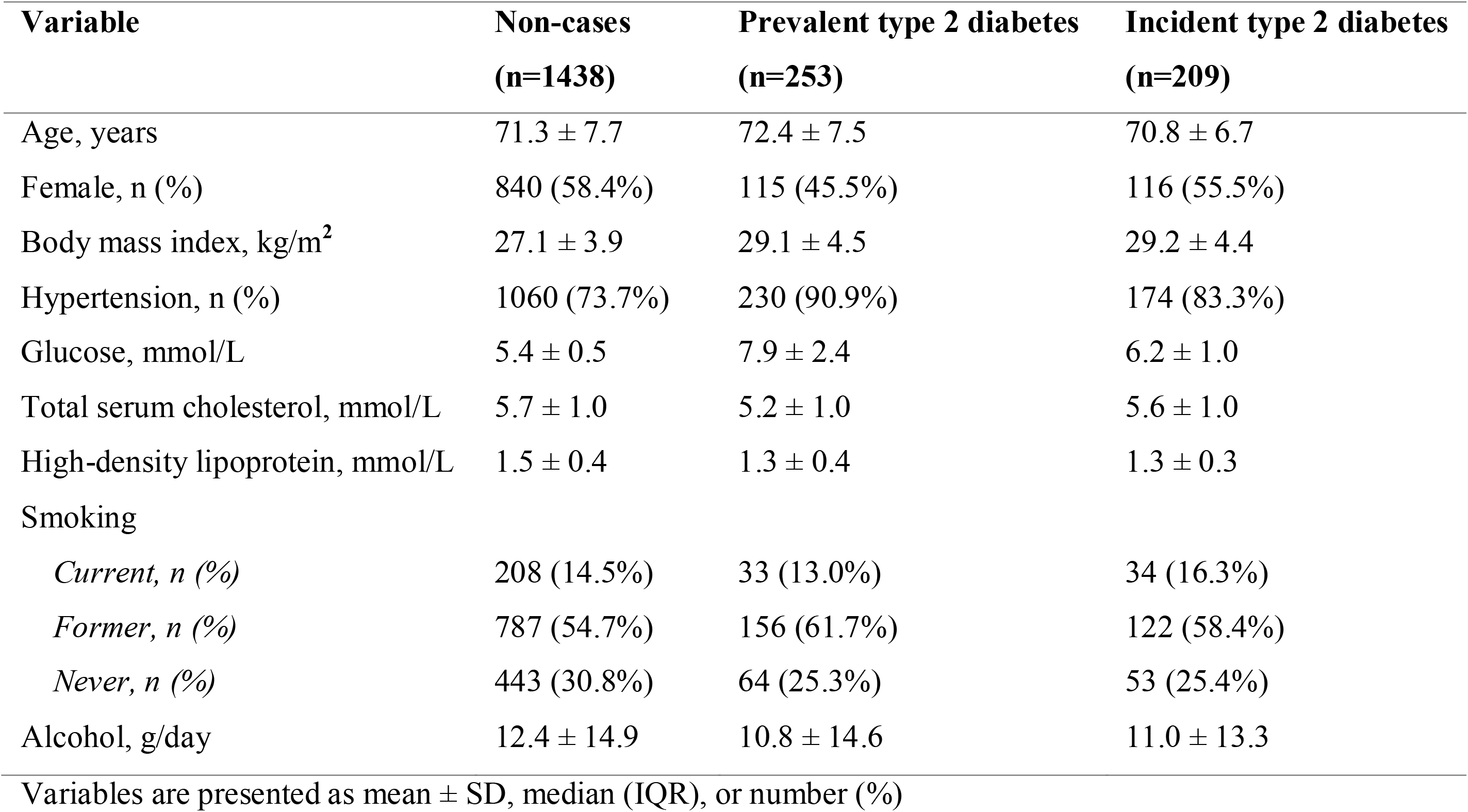
Baseline characteristics of participants of the Rotterdam Study.

| Variable | Non-cases<br>(n=1438) | Prevalent type 2 diabetes<br>(n=253) | Incident type 2 diabetes<br>(n=209) |
| --- | --- | --- | --- |
| Age, years | 71.3 ± 7.7 | 72.4 ± 7.5 | 70.8 ± 6.7 |
| Female, n (%) | 840 (58.4%) | 115 (45.5%) | 116 (55.5%) |
| Body mass index, kg/m<sup>2</sup> | 27.1 ± 3.9 | 29.1 ± 4.5 | 29.2 ± 4.4 |
| Hypertension, n (%) | 1060 (73.7%) | 230 (90.9%) | 174 (83.3%) |
| Glucose, mmol/L | 5.4 ± 0.5 | 7.9 ± 2.4 | 6.2 ± 1.0 |
| Total serum cholesterol, mmol/L | 5.7 ± 1.0 | 5.2 ± 1.0 | 5.6 ± 1.0 |
| High-density lipoprotein, mmol/L | 1.5 ± 0.4 | 1.3 ± 0.4 | 1.3 ± 0.3 |
| Smoking |  |  |  |
| *Current, n (%)* | 208 (14.5%) | 33 (13.0%) | 34 (16.3%) |
| *Former, n (%)* | 787 (54.7%) | 156 (61.7%) | 122 (58.4%) |
| *Never, n (%)* | 443 (30.8%) | 64 (25.3%) | 53 (25.4%) |
| Alcohol, g/day | 12.4 ± 14.9 | 10.8 ± 14.6 | 11.0 ± 13.3 |

Variables are presented as mean ± SD, median (IQR), or number (%)

**Table 2.**
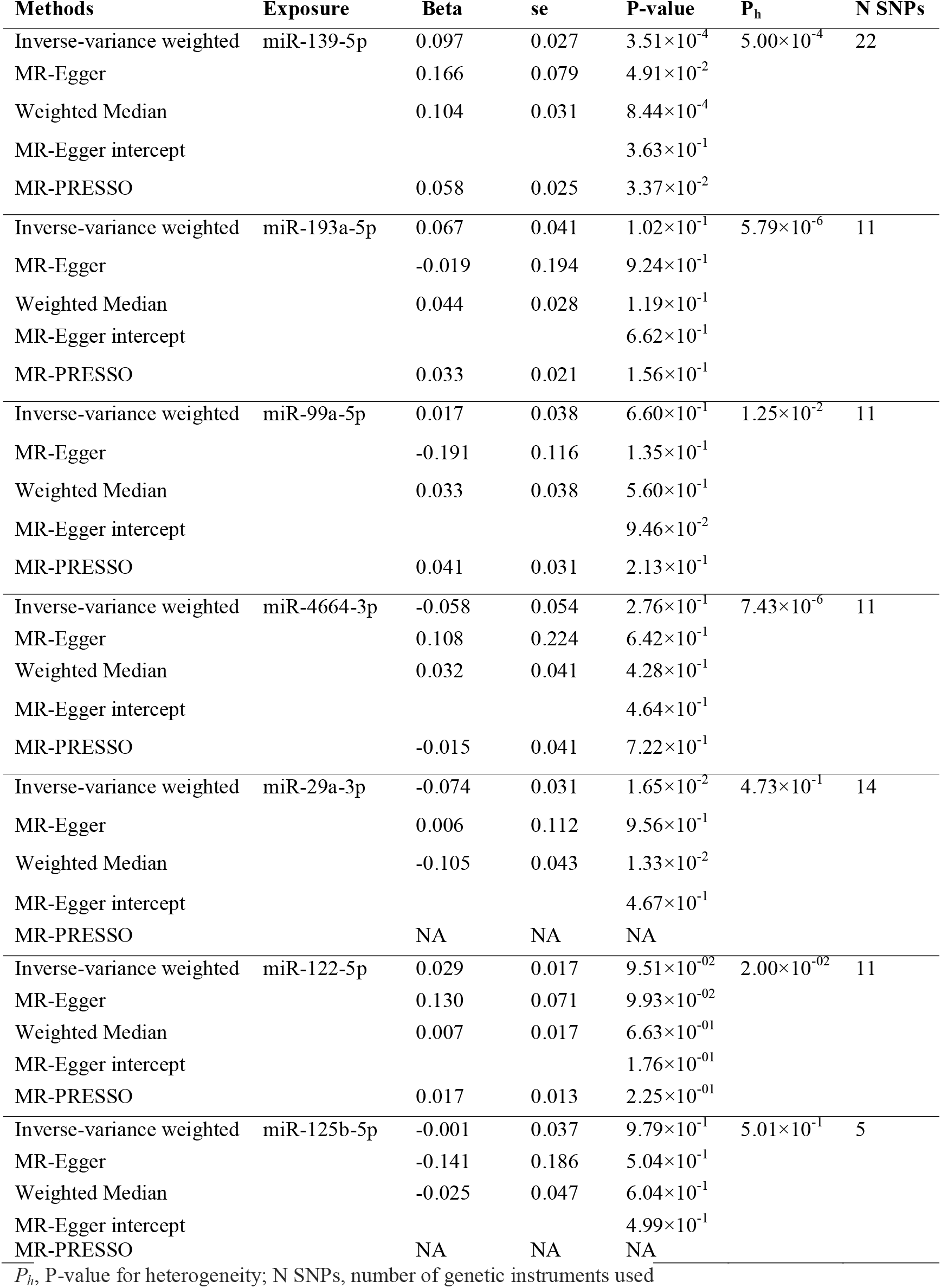
The results of MR analysis between miRNAs associated with type 2 diabetes.

| Methods | Exposure | Beta | se | P-value | P<sub>h</sub> | N SNPs |
| --- | --- | --- | --- | --- | --- | --- |
| Inverse-variance weighted | miR-139-5p | 0.097 | 0.027 | $3.51 \times 10^{-4}$ | $5.00 \times 10^{-4}$ | 22 |
| MR-Egger | | 0.166 | 0.079 | $4.91 \times 10^{-2}$ | | |
| Weighted Median | | 0.104 | 0.031 | $8.44 \times 10^{-4}$ | | |
| MR-Egger intercept | | | | $3.63 \times 10^{-1}$ | | |
| MR-PRESSO | | 0.058 | 0.025 | $3.37 \times 10^{-2}$ | | |
| Inverse-variance weighted | miR-193a-5p | 0.067 | 0.041 | $1.02 \times 10^{-1}$ | $5.79 \times 10^{-6}$ | 11 |
| MR-Egger | | -0.019 | 0.194 | $9.24 \times 10^{-1}$ | | |
| Weighted Median | | 0.044 | 0.028 | $1.19 \times 10^{-1}$ | | |
| MR-Egger intercept | | | | $6.62 \times 10^{-1}$ | | |
| MR-PRESSO | | 0.033 | 0.021 | $1.56 \times 10^{-1}$ | | |
| Inverse-variance weighted | miR-99a-5p | 0.017 | 0.038 | $6.60 \times 10^{-1}$ | $1.25 \times 10^{-2}$ | 11 |
| MR-Egger | | -0.191 | 0.116 | $1.35 \times 10^{-1}$ | | |
| Weighted Median | | 0.033 | 0.038 | $5.60 \times 10^{-1}$ | | |
| MR-Egger intercept | | | | $9.46 \times 10^{-2}$ | | |
| MR-PRESSO | | 0.041 | 0.031 | $2.13 \times 10^{-1}$ | | |
| Inverse-variance weighted | miR-4664-3p | -0.058 | 0.054 | $2.76 \times 10^{-1}$ | $7.43 \times 10^{-6}$ | 11 |
| MR-Egger | | 0.108 | 0.224 | $6.42 \times 10^{-1}$ | | |
| Weighted Median | | 0.032 | 0.041 | $4.28 \times 10^{-1}$ | | |
| MR-Egger intercept | | | | $4.64 \times 10^{-1}$ | | |
| MR-PRESSO | | -0.015 | 0.041 | $7.22 \times 10^{-1}$ | | |
| Inverse-variance weighted | miR-29a-3p | -0.074 | 0.031 | $1.65 \times 10^{-2}$ | $4.73 \times 10^{-1}$ | 14 |
| MR-Egger | | 0.006 | 0.112 | $9.56 \times 10^{-1}$ | | |
| Weighted Median | | -0.105 | 0.043 | $1.33 \times 10^{-2}$ | | |
| MR-Egger intercept | | | | $4.67 \times 10^{-1}$ | | |
| MR-PRESSO |  | NA | NA | NA |  |  |
| Inverse-variance weighted | miR-122-5p | 0.029 | 0.017 | $9.51 \times 10^{-02}$ | $2.00 \times 10^{-02}$ | 11 |
| MR-Egger | | 0.130 | 0.071 | $9.93 \times 10^{-02}$ | | |
| Weighted Median | | 0.007 | 0.017 | $6.63 \times 10^{-01}$ | | |
| MR-Egger intercept | | | | $1.76 \times 10^{-01}$ | | |
| MR-PRESSO | | 0.017 | 0.013 | $2.25 \times 10^{-01}$ | | |
| Inverse-variance weighted | miR-125b-5p | -0.001 | 0.037 | $9.79 \times 10^{-1}$ | $5.01 \times 10^{-1}$ | 5 |
| MR-Egger | | -0.141 | 0.186 | $5.04 \times 10^{-1}$ | | |
| Weighted Median | | -0.025 | 0.047 | $6.04 \times 10^{-1}$ | | |
| MR-Egger intercept | | | | $4.99 \times 10^{-1}$ | | |
| MR-PRESSO |  | NA | NA | NA |  |  |

P<sub>h</sub>, P-value for heterogeneity; N SNPs, number of genetic instruments used

**Figure 2.**
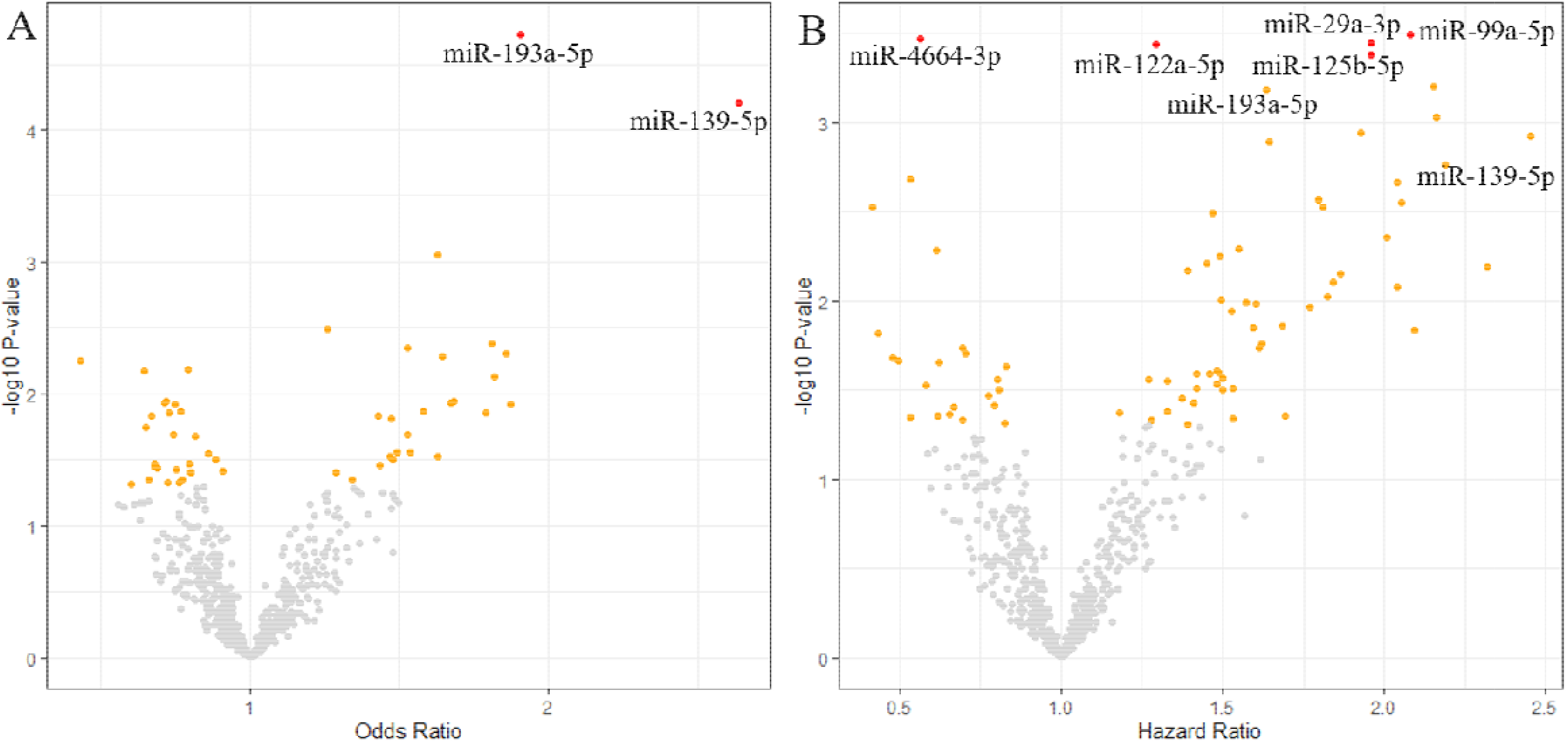
Volcano plots of the effect size of the association between plasma miRNA levels and type 2 diabetes. These plots show the odds ratios of plasma miRNA levels and prevalent diabetes (A) and hazard ratios of plasma miRNA levels and incident diabetes (B). The colors of the dots indicate the significance level: gray, non-significant; orange, nominally associated (P-value<0·05); red, significantly associated (FDR<0·05). Identified miRNAs with at least nominal association are labeled in both plots.

To assess the impact of relative expression of the identified miRNAs on diabetes risk, we categorized the expression values of each miRNA into low and high categories based on the median value. We found that high expression levels of miR-99a-5p (median normalized value >8.12), miR-29a-3p (median normalized value >10.21), miR-125b-5p (median normalized value >8.20), miR-139-5p (median normalized value >8.82) and miR-193a-5p (median normalized value >8.12) were also significantly associated with higher cumulative hazard of diabetes (**Figure 3**).

**Figure 3.**
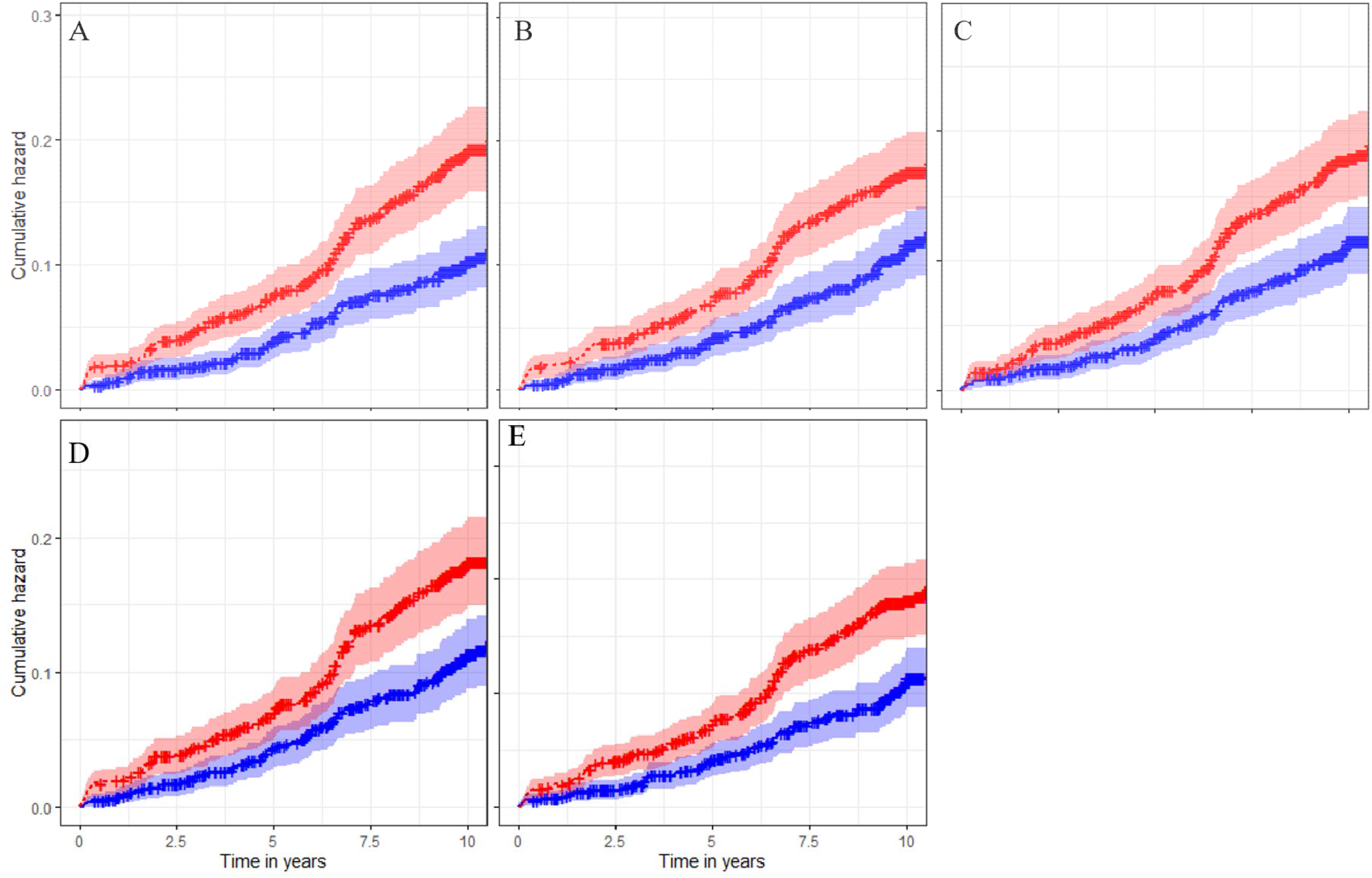
Significant cumulative hazard curves of log2 CPM expression of miRNAs and incident type 2 diabetes. The red curves indicate a high expression level and the blue curves indicate a low expression level based on the log2 median values of individual miRNAs. Y-axis indicates the cumulative hazard. The X-axis indicates the time scale in years. **A**. Cumulative hazard on incident diabetes between high and low expression levels of miR-99a-5p. **B**. Cumulative hazard on incident diabetes between high and low expression levels of miR-29a-3p. **C**. Cumulative hazard on incident diabetes between high and low expression levels of miR-125b-5p. **D**. Cumulative hazard on incident diabetes between high and low expression levels of miR-139-5p. **E**. Cumulative hazard on incident diabetes between high and low expression levels of miR-193a-5p.

Next, we assessed the causal relationship between the identified miRNAs and type 2 diabetes. Running GWAS in the Rotterdam Study (n=1687), we obtained genetic instruments for the seven identified miRNAs and performed two-sample MR. The results of the MR studies are presented in **Table 2**. MR analysis supported a causal association between miR-139-5p and type 2 diabetes (MR-IVW effect estimate 0.10; IVW p=3.51×10^−4^). We found significant heterogeneity for miR-139-5p (p_h_=5.00×10^−4^), however, there was no evidence of directional pleiotropy (MR Egger intercept p=0.36). The IVW effect estimate was in the same direction with those from WM and MR-Egger methods (**Table 2**; **Supplementary figures 2-4**).

To address the potential role of miR-139-5p in regulating metabolic pathways, we tested if the 22 genetic variants, that act in *trans* on the expression levels of miR-139-5p, have reported to also change the expression levels of any metabolites using the previous GWAS on plasma metabolites levels (20). Out of the 22 genetic variants, 21 were associated with at least one metabolite (P<0.05). Over 38% of the SNP-metabolite sets came from phosphatidylcholines, followed by 17% from amino acids (**Figure 4**). A large proportion of the SNP-phosphatidylcholines sets is due to the association of five SNPs located on the Chr9q34.2 locus (rs687289, rs558240, rs176694, rs11244083, rs592514). Additionally, we tested if the 22 genetic variants related to miR-139-5p have been associated to change the expression of genes using the previous GWAS (15). Genes of which their expression levels are strongly regulated by one of the 22 variants are *MED22* (Chr9q34.2), *AP4E1* (Chr15q21.2), *SURF1* (Chr9q34.2), and *SURF6* (Chr9q34.2). The full set of genes can be found in **Supplementary Table 4**. Notably, the aforementioned SNPs related to the phosphatidylcholines are also associated with the expression level of *ABO* on the Chr9q34.2 locus.

**Figure 4.**
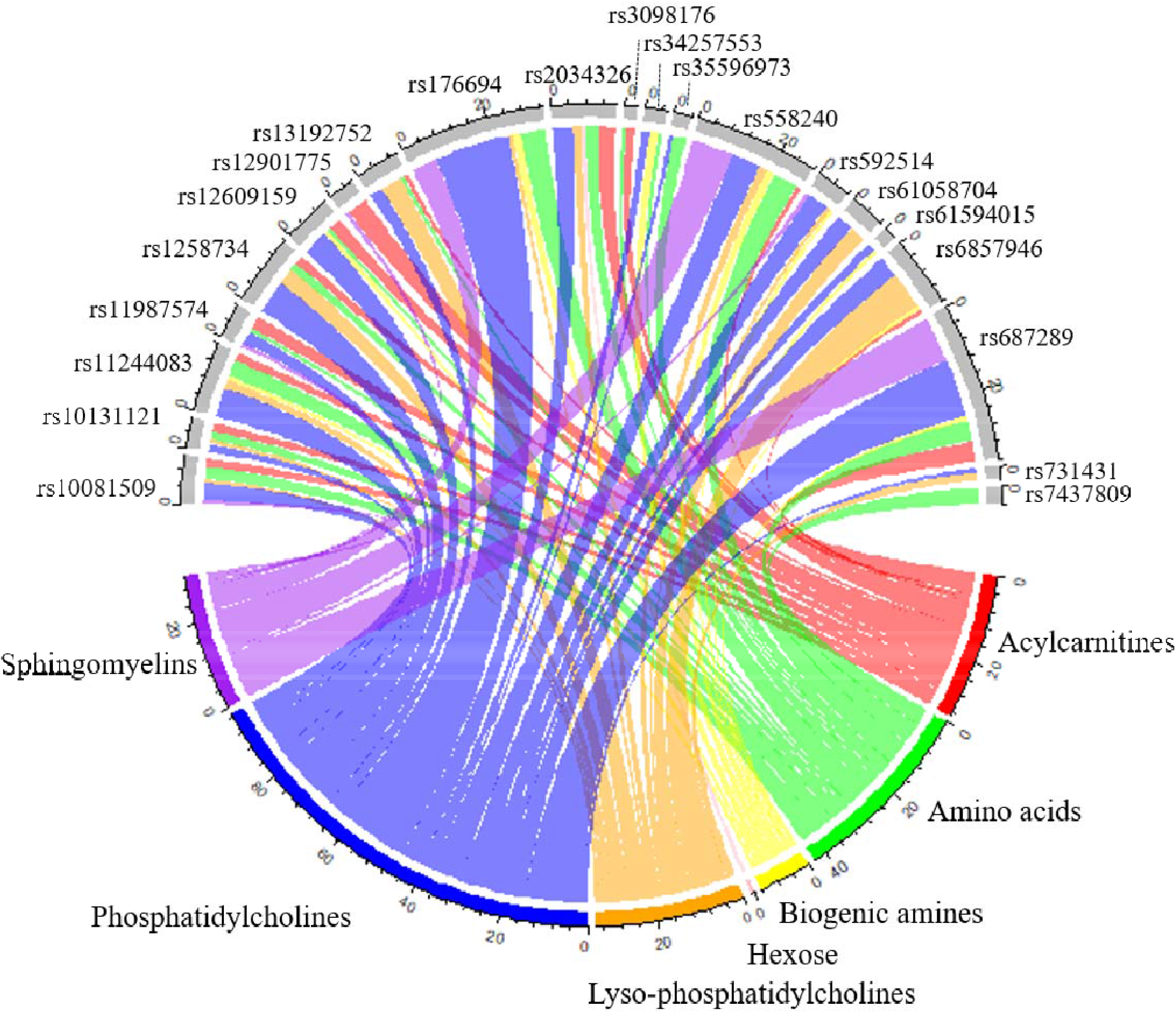
Chord diagram illustrating the link between miR-139-5p SNPs and metabolites. Each color presents a class of metabolite (e.g. sphingomyelins, phosphatidylcholines, lyso-phosphatidylcholines, hexose, biogenic amines, amino acids and acylcarnitines). Each line presents an association between SNP and metabolite

Given that miRNAs control disease risk through regulating the expression of their target genes, we extracted SNPs in 214 target genes of miR-139-5p and tested their association with type 2 diabetes and fasting glucose using summary statistics from recent GWASs (5, 19). We found 68 putative target genes and 5 validated target genes (incl. *BLC2, NR5A2, NFKB1, OIP5, MCL1*) to be associated with type 2 diabetes (FDR<0.05). In addition, we found 13 putative target genes and 2 validated target genes (incl. *IGF1R, NOTCH1*) to be associated with fasting glucose (**Supplementary Table 5**). According to GTEx v8 (21), the associated target genes of miR-139-5p are most specific to be downregulated in pancreatic, kidney, heart and liver tissue. The expression of these target genes across different tissues was obtained using FUMA (21) and can be found in **Supplementary figures 5-6**.

## Discussion

In this study, we characterized 591 well-expressed miRNAs to identify associations with prevalent and incident type 2 diabetes in a population-based cohort. Using an agnostic approach, we found higher plasma levels of miR-139-5p and miR-193a-5p in diabetes patients at the baseline. Moreover, four miRNAs (miR-99a-5p, miR-29a-3p, miR-122-5p and miR-125b-5p) were positively associated with incident diabetes during follow-up. Notable, we found miR-4664-3p to be associated with incident diabetes in an opposite direction, suggesting a protective effect of this miRNA. MR analysis confirmed a causal relationship between the plasma level of miR-139-5p and the risk of type 2 diabetes. Our post-hoc analysis further provided insight into the underlying molecular pathways that could link miR-139-5p to the pathophysiology of type 2 diabetes.

The link between miRNAs and type 2 diabetes has gained increasing attention in recent years (22-24). In particular, population-based prospective cohort studies with long follow-up time are needed to detect differentially expressed miRNAs at different stages of the disease. The observed differences in miRNA profiles between a disease versus a healthy state are a result of tissue-specific processes occurring at different stages of the disease. Therefore, miRNAs in the associations with prevalent and incident type 2 diabetes do not necessarily need to be consistent to serve as disease biomarkers. Out of the seven identified miRNAs in our study, miR-122-5p has been well-studied miRNA concerning diabetes and liver diseases. For example, a previous study (25) reported an association between miR-122-5p with type 2 diabetes, impaired fasting glucose, and HbA1c despite a small number of cases (n=24). Another study (24) found a higher risk for metabolic syndrome and diabetes over 15 years and additionally reported a strong correlation between miR-122, and lipid levels (triglycerides, LDL and HDL). Furthermore, we recently demonstrated that miR-122-5p is linked to fatty liver disease, which is a common problem in patients with diabetes (7).

In this study, we used a genome-wide sequencing-based miRNA profiling method to investigate the causal association between plasma-derived miRNA expression and type 2 diabetes. The consistency of findings between regression analysis in the observational data and MR analysis supports a causal role for miR-139-5p in developing type 2 diabetes. MR exploits the random allocation of genetic variants during conception that is unlikely to be subject to reverse causation and confounding, therefore is comparable to randomly allocated intervention in a clinical trial (3). Different sensitivity analyses for MR has also supported a robust estimate of causal effects. Collectively, our findings indicate that higher levels of miR-139-5p in plasma increase the risk of diabetes, which in addition to its potential as biomarker, might have therapeutic potential that warrant further investigation.

The increasing interest in discovering miRNAs for complex traits such as diabetes, has led to better miRNA quantification platforms. In particular, the EdgeSeq array, which has been used in the current study, is a high-throughput technology that is reported to be more accurate, sensitive and specific when compared to a traditional RT-qPCR method (26). Although this EdgeSeq method is still little used in epidemiological studies, it has been shown to detect novel miRNAs with the least bias detection (26) A recent study (27) using the same EdgeSeq array has also researched genetic variants associated with expression levels of miR-139-5p. In the latter study, they focused on different cardio-metabolic phenotypes, but not on diabetes, including lipid and glycemic traits and coronary artery disease. Also, a second study (28) that measured the expression of 750 whole-blood derived miRNAs by RT-qPCR, had identified *cis*-regulatory variants that regulate miR-139-5p. They tested the causal role of miR-139-5p on diabetes but did not find any significant effect. A possible explanation for this includes the differences in miRNA source. Given that whole blood reflects platelet and electrolyte cellular miRNAs, among other things, it may show conflicting expression from extracellular sources, such as plasma.(29) Previously identified SNPs for miR-139-5p were all regulating in *cis* (27, 28), some of which were successfully replicated in our GWAS (p<0.05). We were able to identify *trans-*regulatory variants for miR-139-5p due to the relatively larger sample size in our study compared to the previously mentioned study (27) which used the same miRNA profiling EdgeSeq array. Our study, therefore, has a bigger power to identify genetic instruments for miRNAs for MR analysis.

MiRNA expression is often tissue specific, however, little is known about the tissue-specificity of miR-139-5p in humans. Since miR-139-5p was reported to be embedded within the intron region of *PDE2A* in a sense direction and likely to be co-expressed, we tested whether *PDE2A* was expressed in tissue relevant to diabetes, including pancreas and liver. According to the Human Protein Atlas (https://www.proteinatlas.org/), *PDE2A* is among others expressed in endocrine tissue and mainly in the adrenal gland (30). Moreover, *PDE2A* is reported to be expressed in endothelial cells in liver and pancreas.

In order to test whether miR-139-5p is expressed in tissue relevant to diabetes, such as the pancreas and liver, the host gene (PDE2A) that share the same promoter was used as proxy. Finally, according to FANTOM5(31), miR-139-5p is similar to *PDE2A* (30) enriched in endothelial cells, of which dysfunction is a major mediator of diabetes (32).

As a secondary aim of this study, we investigated downstream effects, including metabolites and target genes to gain better insights into the potential mechanisms miR-139-5p is involved in the pathogenesis of diabetes. We described the association between miR-139-5p and metabolites, of which phosphatidylcholines were the most abundant. Phosphatidylcholines are phospholipids attached to a choline particle and can be found in foods, such as eggs and liver. A recent prospective study (33) has shown a protective effect of phosphatidylcholines intake and diabetes. It is plausible that miR-139-5p plays role in the metabolism of phosphatidylcholines and thus influences the risk of diabetes. Furthermore, we found multiple putative and validated target genes of miR-139-5p that are known for their role in pathways underlying diabetes. For example, *BLC2* is involved in regulating both pro-inflammatory cytokines, glucose metabolism, and pancreatic β-cell homeostasis (34). Another target gene *IGF1R* is a tyrosine kinase receptor that is activated by *IGF-1* and is related to risk for diabetes. This is in line with a previous study (35) that found a positive relation between miR-139-5p expression and the suppression of *IRS1* gene in diabetic rats. *IRS1* plays a major role in transmitting signals from *IGF-1* to downstream cellular pathways such as the MAPK pathway. Moreover, a depressed function of *IRS1* is associated with insulin resistance and pancreatic β-cell function (35).

Strengths of our study include, first, the investigation for well-expressed miRNAs in an agnostic manner in the population level, which could identify novel miRNAs that were not previously recognized for type 2 diabetes. Second, the use of the EdgeSeq array that allowed us to measure the levels of cell-free miRNAs in plasma in a highly specific, sensitive, and reproducible manner. Additionally, we triangulated the evidence integrating cross-sectional, and longitudinal analysis of observational data with MR analysis to examine the causal association of circulatory miRNAs and type 2 diabetes. However, some limitations should be acknowledged when interpreting the results of this study. First, during the baseline examination of this study, traditional parameters such as insulin, HbA1c, HOMA-β, and HOMA-IR were not measured. Therefore, we could not calculate the differences in disease prediction capacity between the identified miRNAs and the abovementioned parameters. Second, glucose and insulin metabolism take place in specific tissues, such as the pancreas and liver, but the analyzed miRNAs in the current study are cell-free in plasma, meaning that the tissue of origin is unknown. As miRNAs are tissue-specific, it would be interesting yet to study the identified miRNAs in relevant tissues and cell types. Moreover, further experimental validation of the identified miRNAs is warranted to confirm their regulatory roles in the pathophysiology of diabetes.

In summary, this study indicates that plasma levels of two miRNAs linked to prevalent diabetes and five miRNAs associated with incident type 2 diabetes. The identified miRNAs are possibly involved in pathways underlying the metabolic disturbance of type 2 diabetes. In particular, elevated levels of miR-139-5p have a causal effect on type 2 diabetes potentially through affecting several important genes and metabolites involved in the disease pathology.

## Data Availability

Requests to access the dataset from qualified researchers trained in human subject confidentiality protocols may be sent to Department of Epidemiology, Erasmus MC University Medical Center.

## Contributors

MG and AD designed the study and oversaw the research. MMJM wrote the first draft of the manuscript. MMJM and RM did the statistical analyses. MG, FA, MAI, MK, and ME provided resources and data. All authors interpreted the data and commented on the draft report. All authors approved the final manuscript.

## References

1. Zampetaki A, Kiechl S, Drozdov I, Willeit P, Mayr U, Prokopi M, et al. Plasma microRNA profiling reveals loss of endothelial miR-126 and other microRNAs in type 2 diabetes. Circ Res. 2010;107(6):810–7.

2. Ortega FJ, Mercader JM, Moreno-Navarrete JM, Rovira O, Guerra E, Esteve E, et al. Profiling of circulating microRNAs reveals common microRNAs linked to type 2 diabetes that change with insulin sensitization. Diabetes Care. 2014;37(5):1375–83.

3. Smith GD, Ebrahim S. ‘Mendelian randomization’: can genetic epidemiology contribute to understanding environmental determinants of disease? Int J Epidemiol. 2003;32(1):1–22.

4. Lotta LA, Pietzner M, Stewart ID, Wittemans LBL, Li C, Bonelli R, et al. A cross-platform approach identifies genetic regulators of human metabolism and health. Nature Genetics. 2021;53(1):54–64.

5. Mahajan A, Taliun D, Thurner M, Robertson NR, Torres JM, Rayner NW, et al. Fine-mapping type 2 diabetes loci to single-variant resolution using high-density imputation and islet-specific epigenome maps. Nat Genet. 2018;50(11):1505–13.

6. Ikram MA, Brusselle G, Ghanbari M, Goedegebure A, Ikram MK, Kavousi M, et al. Objectives, design and main findings until 2020 from the Rotterdam Study. European Journal of Epidemiology. 2020.

7. Zhang X, Mens MMJ, Abozaid YJ, Bos D, Darwish Murad S, de Knegt RJ, et al. Circulatory microRNAs as potential biomarkers for fatty liver disease: the Rotterdam study. Aliment Pharmacol Ther. 2020.

8. World Health O, International Diabetes F. Definition and diagnosis of diabetes mellitus and intermediate hyperglycaemia : report of a WHO/IDF consultation. Geneva: World Health Organization; 2006.

9. van Buuren S, Groothuis-Oudshoorn K. mice: Multivariate Imputation by Chained Equations in R. 2011. 2011;45(3):67.

10. Benjamini Y, Hochberg Y. Controlling the False Discovery Rate - a Practical and Powerful Approach to Multiple Testing. J R Stat Soc B. 1995;57(1):289–300.

11. Bowden J, Del Greco MF, Minelli C, Davey Smith G, Sheehan N, Thompson J. A framework for the investigation of pleiotropy in two-sample summary data Mendelian randomization. Stat Med. 2017;36(11):1783–802.

12. Bowden J, Davey Smith G, Burgess S. Mendelian randomization with invalid instruments: effect estimation and bias detection through Egger regression. Int J Epidemiol. 2015;44(2):512-25.

13. Bowden J, Davey Smith G, Haycock PC, Burgess S. Consistent Estimation in Mendelian Randomization with Some Invalid Instruments Using a Weighted Median Estimator. Genet Epidemiol. 2016;40(4):304–14.

14. Hemani G, Zheng J, Elsworth B, Wade KH, Haberland V, Baird D, et al. The MR-Base platform supports systematic causal inference across the human phenome. Elife. 2018;7.

15. Võsa U, Claringbould A, Westra H-J, Bonder MJ, Deelen P, Zeng B, et al. Unraveling the polygenic architecture of complex traits using blood eQTL metaanalysis. bioRxiv. 2018:447367.

16. Chou C-H, Shrestha S, Yang C-D, Chang N-W, Lin Y-L, Liao K-W, et al. miRTarBase update 2018: a resource for experimentally validated microRNA-target interactions. Nucleic Acids Research. 2017;46(D1):D296–D302.

17. Agarwal V, Bell GW, Nam JW, Bartel DP. Predicting effective microRNA target sites in mammalian mRNAs. Elife. 2015;4.

18. Chen Y, Wang X. miRDB: an online database for prediction of functional microRNA targets. Nucleic Acids Res. 2020;48(D1):D127–D31.

19. Lagou V, Mägi R, Hottenga J-J, Grallert H, Perry JRB, Bouatia-Naji N, et al. Sex-dimorphic genetic effects and novel loci for fasting glucose and insulin variability. Nature Communications. 2021;12(1):24.

20. Lotta LA, Pietzner M, Stewart ID, Wittemans LBL, Li C, Bonelli R, et al. Cross-platform genetic discovery of small molecule products of metabolism and application to clinical outcomes. bioRxiv. 2020:2020.02.03.932541.

21. Watanabe K, Taskesen E, van Bochoven A, Posthuma D. Functional mapping and annotation of genetic associations with FUMA. Nature Communications. 2017;8(1):1826.

22. Jaeger A, Zollinger L, Saely CH, Muendlein A, Evangelakos I, Nasias D, et al. Circulating microRNAs -192 and -194 are associated with the presence and incidence of diabetes mellitus. Sci Rep. 2018;8(1):14274.

23. Jiménez-Lucena R, Camargo A, Alcalá-Diaz JF, Romero-Baldonado C, Luque RM, van Ommen B, et al. A plasma circulating miRNAs profile predicts type 2 diabetes mellitus and prediabetes: from the CORDIOPREV study. Experimental & Molecular Medicine. 2018;50(12):1–12.

24. Willeit P, Skroblin P, Moschen AR, Yin X, Kaudewitz D, Zampetaki A, et al. Circulating MicroRNA-122 Is Associated With the Risk of New-Onset Metabolic Syndrome and Type 2 Diabetes. Diabetes. 2017;66(2):347–57.

25. Mononen N, Lyytikäinen L-P, Seppälä I, Mishra PP, Juonala M, Waldenberger M, et al. Whole blood microRNA levels associate with glycemic status and correlate with target mRNAs in pathways important to type 2 diabetes. Scientific Reports. 2019;9(1):8887.

26. Godoy PM, Barczak AJ, DeHoff P, Srinivasan S, Etheridge A, Galas D, et al. Comparison of Reproducibility, Accuracy, Sensitivity, and Specificity of miRNA Quantification Platforms. Cell Reports. 2019;29(12):4212-22.e5.

27. Nikpay M, Beehler K, Valsesia A, Hager J, Harper ME, Dent R, et al. Genome-wide identification of circulating-miRNA expression quantitative trait loci reveals the role of several miRNAs in the regulation of cardiometabolic phenotypes. Cardiovasc Res. 2019;115(11):1629–45.

28. Huan T, Rong J, Liu C, Zhang X, Tanriverdi K, Joehanes R, et al. Genome-wide identification of microRNA expression quantitative trait loci. Nat Commun. 2015;6:6601.

29. Shah R, Tanriverdi K, Levy D, Larson M, Gerstein M, Mick E, et al. Discordant Expression of Circulating microRNA from Cellular and Extracellular Sources. PLoS One. 2016;11(4):e0153691.

30. Uhlen M, Fagerberg L, Hallstrom BM, Lindskog C, Oksvold P, Mardinoglu A, et al. Proteomics. Tissue-based map of the human proteome. Science. 2015;347(6220):1260419.

31. de Rie D, Abugessaisa I, Alam T, Arner E, Arner P, Ashoor H, et al. An integrated expression atlas of miRNAs and their promoters in human and mouse. Nature Biotechnology. 2017;35(9):872–8.

32. Kaur R, Kaur M, Singh J. Endothelial dysfunction and platelet hyperactivity in type 2 diabetes mellitus: molecular insights and therapeutic strategies. Cardiovasc Diabetol. 2018;17(1):121.

33. Virtanen JK, Tuomainen T-P, Voutilainen S. Dietary intake of choline and phosphatidylcholine and risk of type 2 diabetes in men: The Kuopio Ischaemic Heart Disease Risk Factor Study. European Journal of Nutrition. 2020;59(8):3857–61.

34. Gurzov EN, Eizirik DL. Bcl-2 proteins in diabetes: mitochondrial pathways of beta-cell death and dysfunction. Trends Cell Biol. 2011;21(7):424–31.

35. Li J, Su L, Gong YY, Ding ML, Hong SB, Yu S, et al. Downregulation of miR-139-5p contributes to the antiapoptotic effect of liraglutide on the diabetic rat pancreas and INS-1 cells by targeting IRS1. PLoS One. 2017;12(3):e0173576.

