## Supplementary figures for "MiR-139-5p is a causal biomarker for type 2 diabetes; Results from genome-wide microRNA profiling and Mendelian randomization analysis in a population-based study"

**Supplementary material**

Supplementary Figure 1. Forest plot of the association between miRNA expression and prevalent and incident diabetes stratified by cohort

Supplementary Figure 2. The effect for individual genetic instruments for miR-139-5p and type 2 diabetes

Supplementary Figure 3. Scatter plot of different MR analysis

Supplementary Figure 4. Leave-one-out plot of MR analysis

Supplementary Figure 5. Differential expression of miR-139-5p target genes across tissues

Supplementary Figure 6. Heatmap of expressed miR-139-5p target genes across tissues

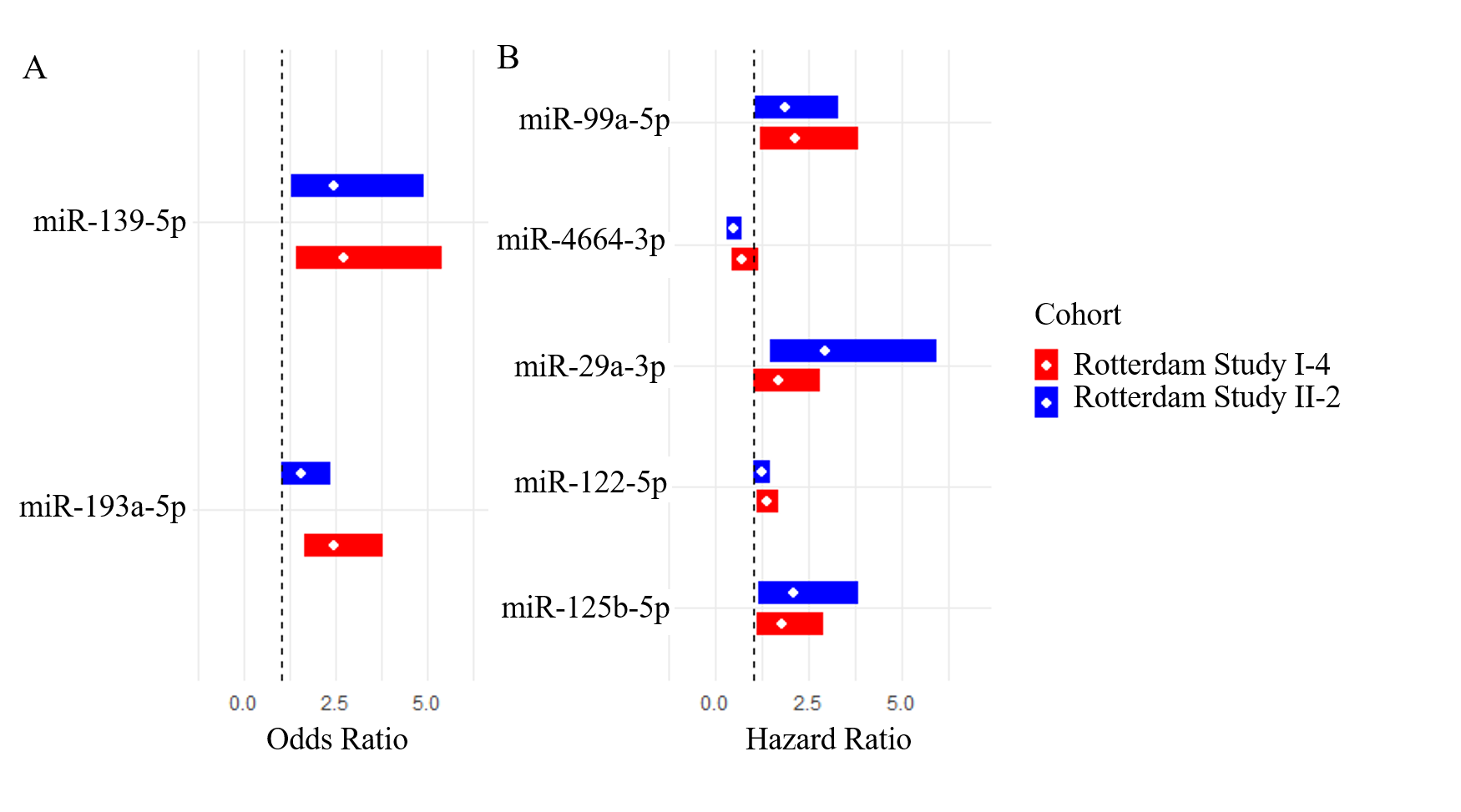

| Rotterdam Study-I-4 (n=912) | | | | Rotterdam Study-II-2 (n=988) | | | |
| --- | --- | --- | --- | --- | --- | --- | --- |
| Prevalent diabetes (n=126) | | | | **Prevalent diabetes (n=127)** | | | |
| miRNA | OR | 95%CI | P value | miRNA | OR | 95%CI | P value |
| miR-139-5p | 2.71 | 1.40-5.41 | 3.89×10^-3^ | miR-139-5p | 2.47 | 1.28-4.91 | 8.25×10^-3^ |
| miR-193a-5p | 2.47 | 1.62-3.79 | 2.82×10^-5^ | miR-193a-5p | 1.54 | 1.01-2.35 | 4.59×10^-2^ |
| Incident diabetes (n=103) | | | | **Incident diabetes (n=106)** | | | |
| miRNA | HR | 95%CI | P value | miRNA | HR | 95%CI | P value |
| miR-99a-5p | 2.15 | 1.20-3.85 | 9.64×10^-3^ | miR-99a-5p | 1.88 | 1.08-3.29 | 2.62×10^-2^ |
| miR-4664-3p | 0.69 | 0.43-1.13 | 1.43×10^-1^ | miR-4664-3p | 0.47 | 0.31-0.72 | 6.04×10^-4^ |
| miR-29a-3p | 1.69 | 1.02-2.80 | 4.04×10^-2^ | miR-29a-3p | 2.94 | 1.45-5.94 | 2.71×10^-3^ |
| miR-122-5p | 1.36 | 1.09-1.69 | 6.23×10^-3^ | miR-122-5p | 1.22 | 1.01-1.47 | 4.39×10^-2^ |
| miR-125b-5p | 1.78 | 1.09-2.90 | 2.20×10^-2^ | miR-125b-5p | 2.08 | 1.13-3.82 | 1.82×10^-2^ |

**Supplementary Figure 1**. **Forest plot of the association between miRNA expression and prevalent and incident diabetes stratified by cohort.** Results are displayed as the odds ratio for prevalent diabetes (A), and hazard ratio for incident diabetes (B) with 95% confidence interval. In red displayed the results of Rotterdam Study I-4. In blue displayed the results of Rotterdam Study II-2. The table presents the odds ratio or hazard ratio with 95%CI and P-value.

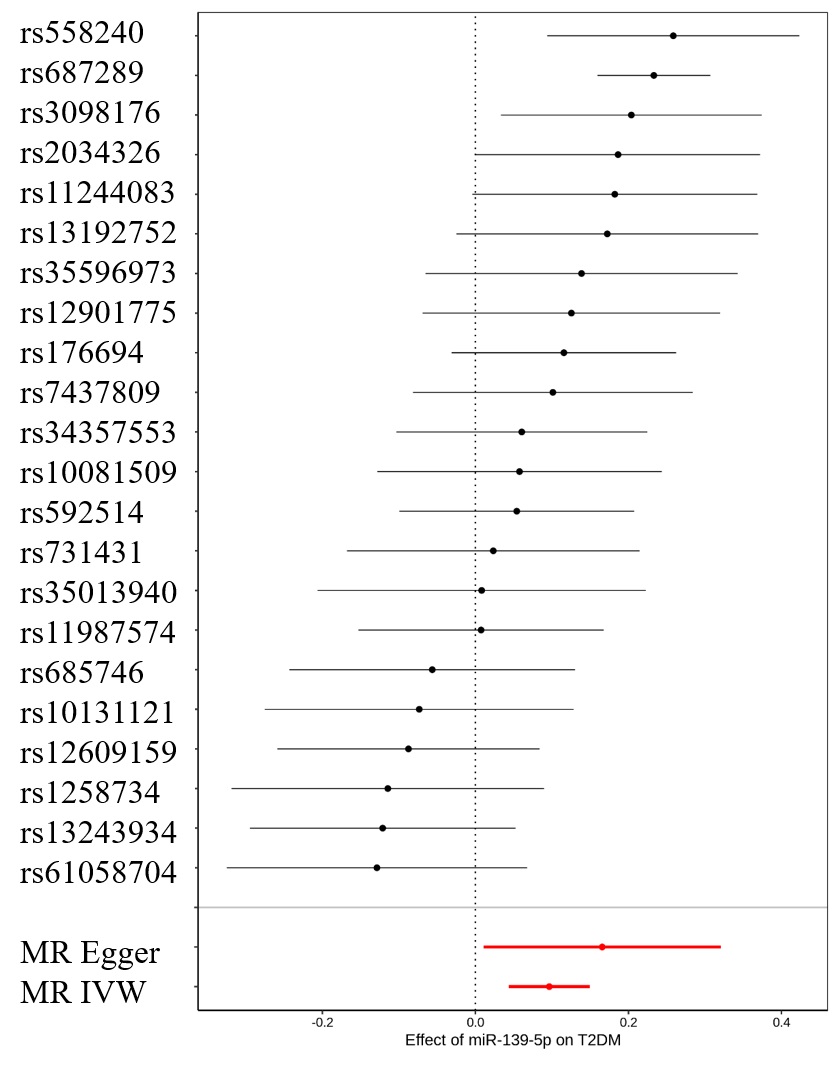

**Supplementary Figure 2**. **The effect for individual genetic instruments for miR-139-5p and type 2 diabetes.** Results are displayed as Beta estimates with 95% CIs. The x-axis presents the effect for each of the individual genetic instrumental variables and type 2 diabetes. In red are displayed the results of MR Egger and MR IVW.

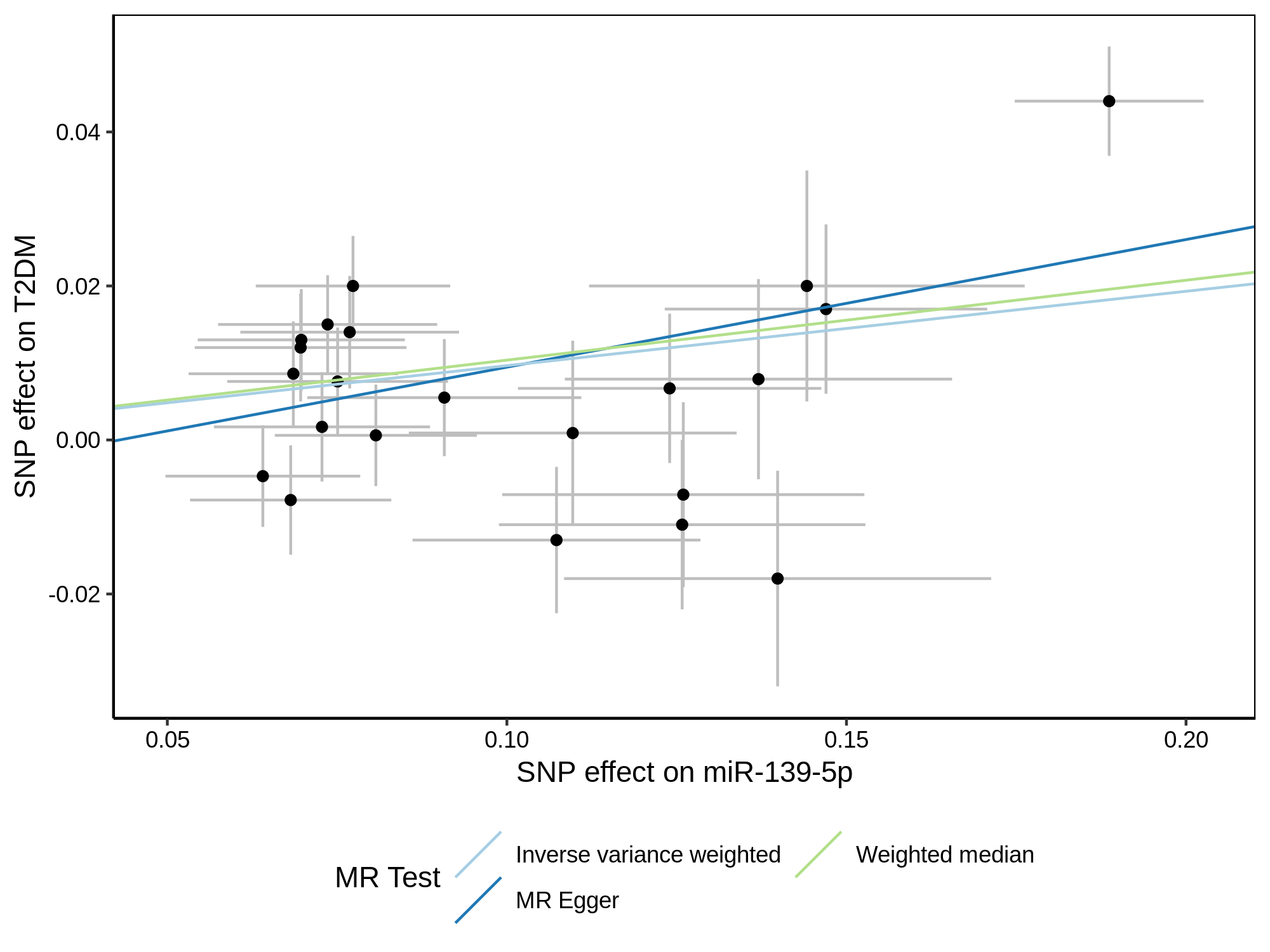

**Supplementary Figure 3. Scatter plot of different MR analysis.** Each dot represents an individual genetic instrument with 95%CI. The x-axis presents the individual effect of each genetic variant on miR-139-5p and the y-axis presents the effect on type 2 diabetes. Each color presents a different MR test; light blue MR Inverse variance weighted; dark blue MR Egger; green MR Weighted median.

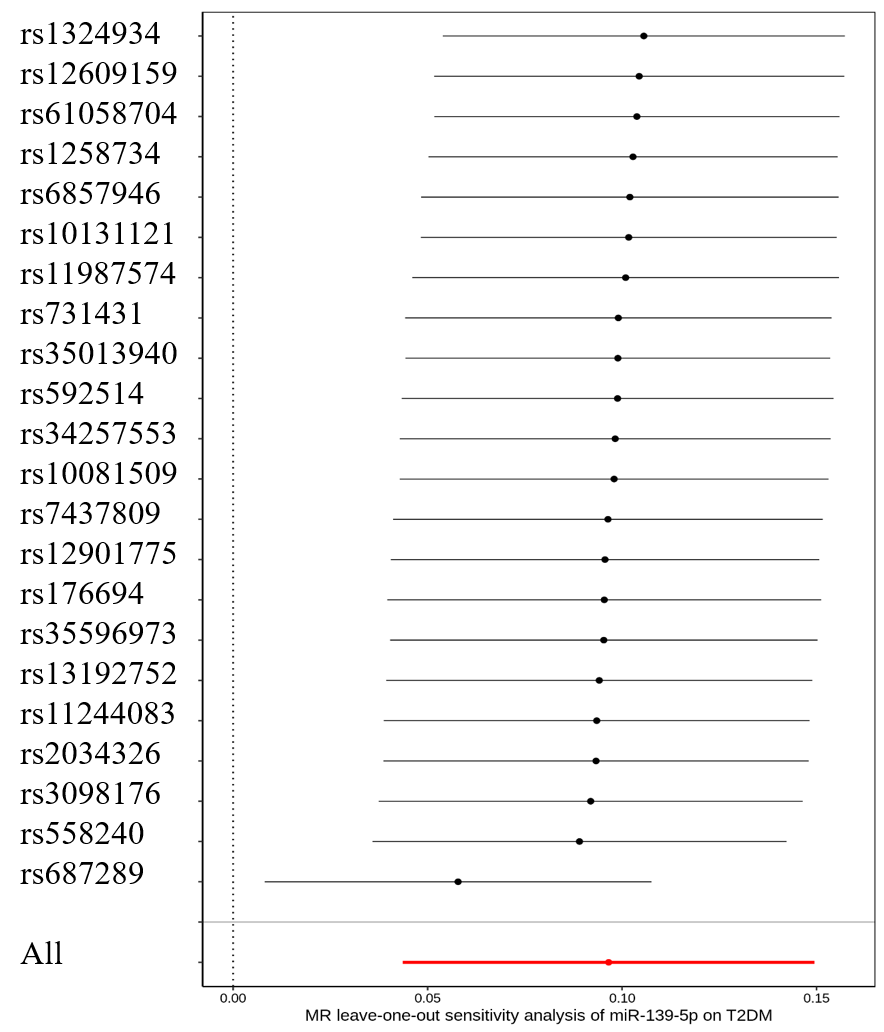

**Supplementary Figure 4. Leave-one-out plot of MR analysis**. This sensitivity analysis of MR analysis (using IVW) excludes a particular genetic instrument. Red presents the overall effect.

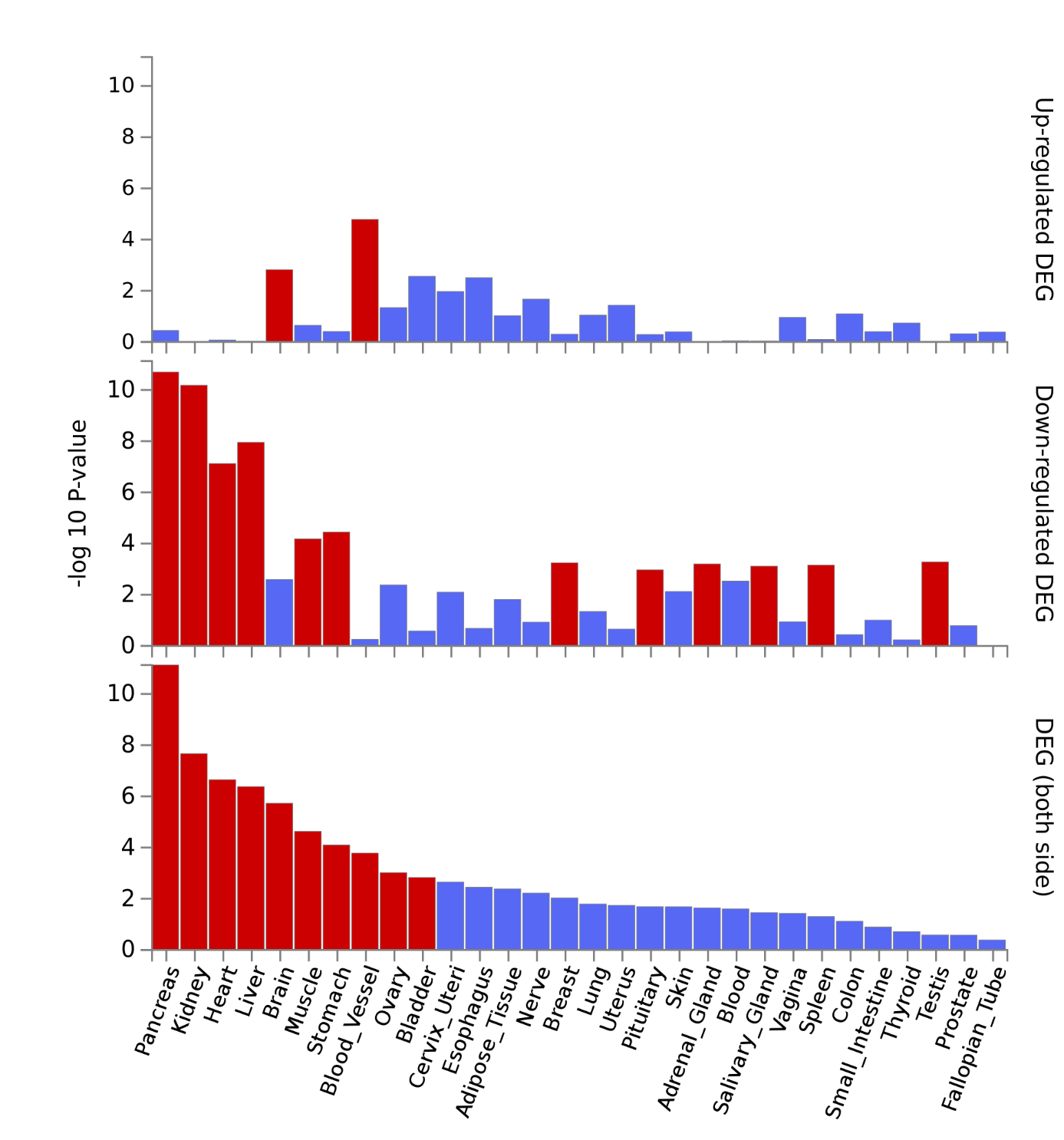

**Supplementary Figure 5. Differential expression of miR-139-5p target genes across tissues.** The top graph presents upregulated genes per tissue, the middle graph down-regulated genes, and the bottom graph overall differential expressed genes. Red color presents significant differences.

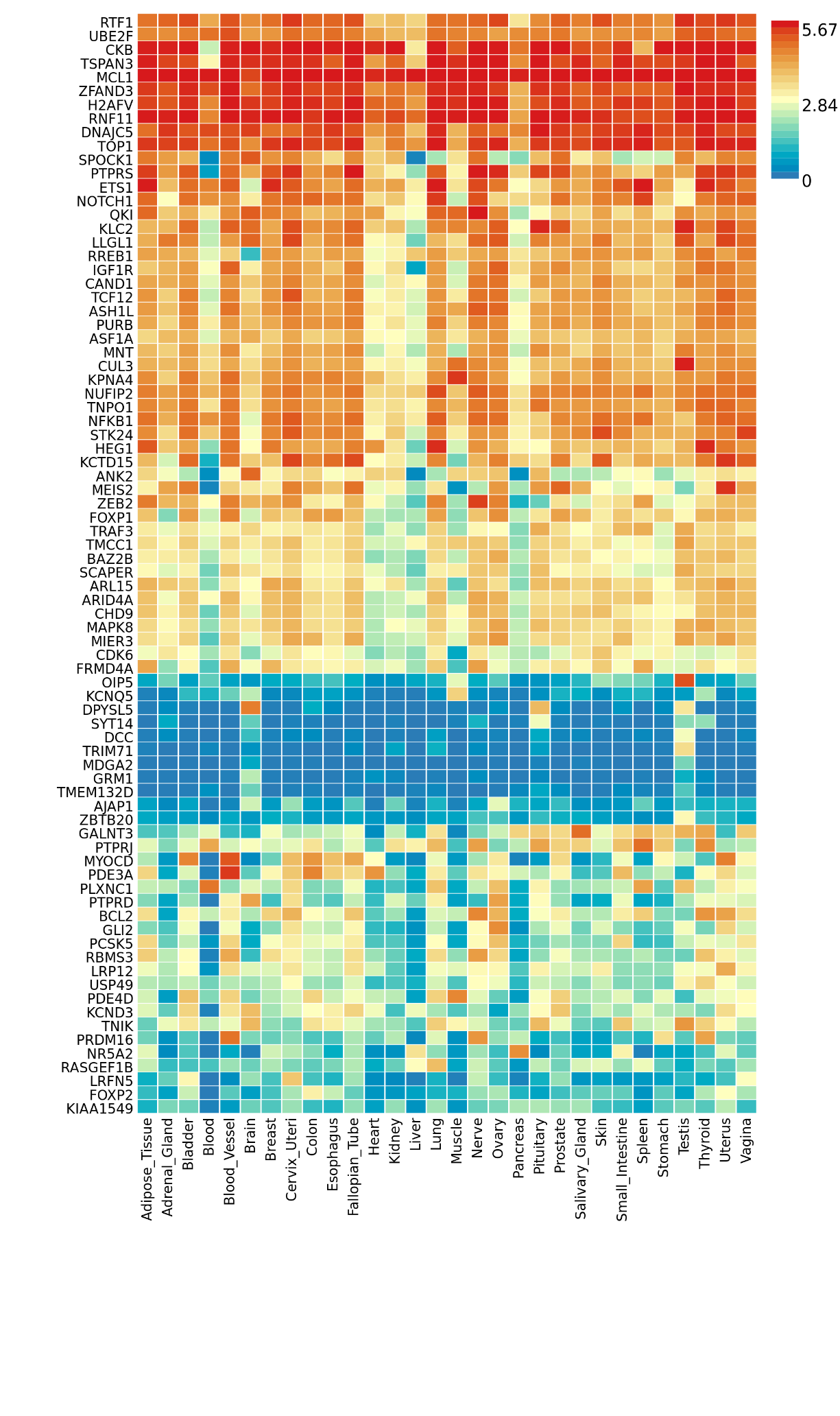

**Supplementary Figure 6. Heatmap of expressed miR-139-5p target genes across tissues.** Red indicates a relatively high expression whereas blue indicates a relatively low expression.
